# Development of a privacy preserving large language model for automated data extraction from thyroid cancer pathology reports

**DOI:** 10.1101/2023.11.08.23298252

**Authors:** Denise T Lee, Akhil Vaid, Kartikeya M Menon, Robert Freeman, David S Matteson, Michael P Marin, Girish N Nadkarni

## Abstract

**Background:** Popularized by ChatGPT, large language models (LLM) are poised to transform the scalability of clinical natural language processing (NLP) downstream tasks such as medical question answering (MQA) and may enhance the ability to rapidly and accurately extract key information from clinical narrative reports. However, the use of LLMs in the healthcare setting is limited by cost, computing power and concern for patient privacy. In this study we evaluate the extraction performance of a privacy preserving LLM for automated MQA from surgical pathology reports.

**Methods:** 84 thyroid cancer surgical pathology reports were assessed by two independent reviewers and the open-source FastChat-T5 3B-parameter LLM using institutional computing resources. Longer text reports were converted to embeddings. 12 medical questions for staging and recurrence risk data extraction were formulated and answered for each report. Time to respond and concordance of answers were evaluated.

**Results:** Out of a total of 1008 questions answered, reviewers 1 and 2 had an average concordance rate of responses of 99.1% (SD: 1.0%). The LLM was concordant with reviewers 1 and 2 at an overall average rate of 88.86% (SD: 7.02%) and 89.56% (SD: 7.20%). The overall time to review and answer questions for all reports was 206.9, 124.04 and 19.56 minutes for Reviewers 1, 2 and LLM, respectively.

**Conclusion:** A privacy preserving LLM may be used for MQA with considerable time-saving and an acceptable accuracy in responses. Prompt engineering and fine tuning may further augment automated data extraction from clinical narratives for the provision of real-time, essential clinical insights.

## Introduction

Surgical pathology reports contain narrative data essential for comprehensive cancer surveillance databases and real-time understanding of staging, recurrence risk, clinical trial eligibility and individual treatment options. However, the large-scale extraction of key surgical oncologic insights contained within unstructured free pathology text is limited by the need for labor-intensive, manual review or error-prone natural language processing (NLP) based on statistical or rule-based approaches (1, 2). Large language models (LLM) power a new generation of natural language processing (NLP) whereby deep neural networks are trained on human language deconstructed into vectorized embeddings that depict linguistic relationships in a numerical format appropriate for easy analysis (3). Popularized by ChatGPT and its user-friendly question and answer interface, LLMs are poised to transform the scalability of clinical NLP downstream tasks such as medical question answering (MQA) and may enhance the ability to rapidly and accurately extract key information from surgical pathology reports (4).

However, ethical, privacy and regulatory constraints preclude the transfer of protected health information (PHI) across the public domain through widely used LLM services (ChatGPT, Bard) that can generate automated responses for MQA. Furthermore, proprietary LLMs may be both expensive, and subject to unpredictable changes in performance. To address these issues, we develop a framework to utilize a privacy-preserving, local LLM for extracting key staging and recurrence risk information from thyroid surgical pathology reports. Although we utilize this for a single use case, our work serves as a novel paradigm to enable individual medical centers to utilize LLM technology for clinical NLP tasks in a privacy protecting manner (**Figure 1**).

**Figure 1.**
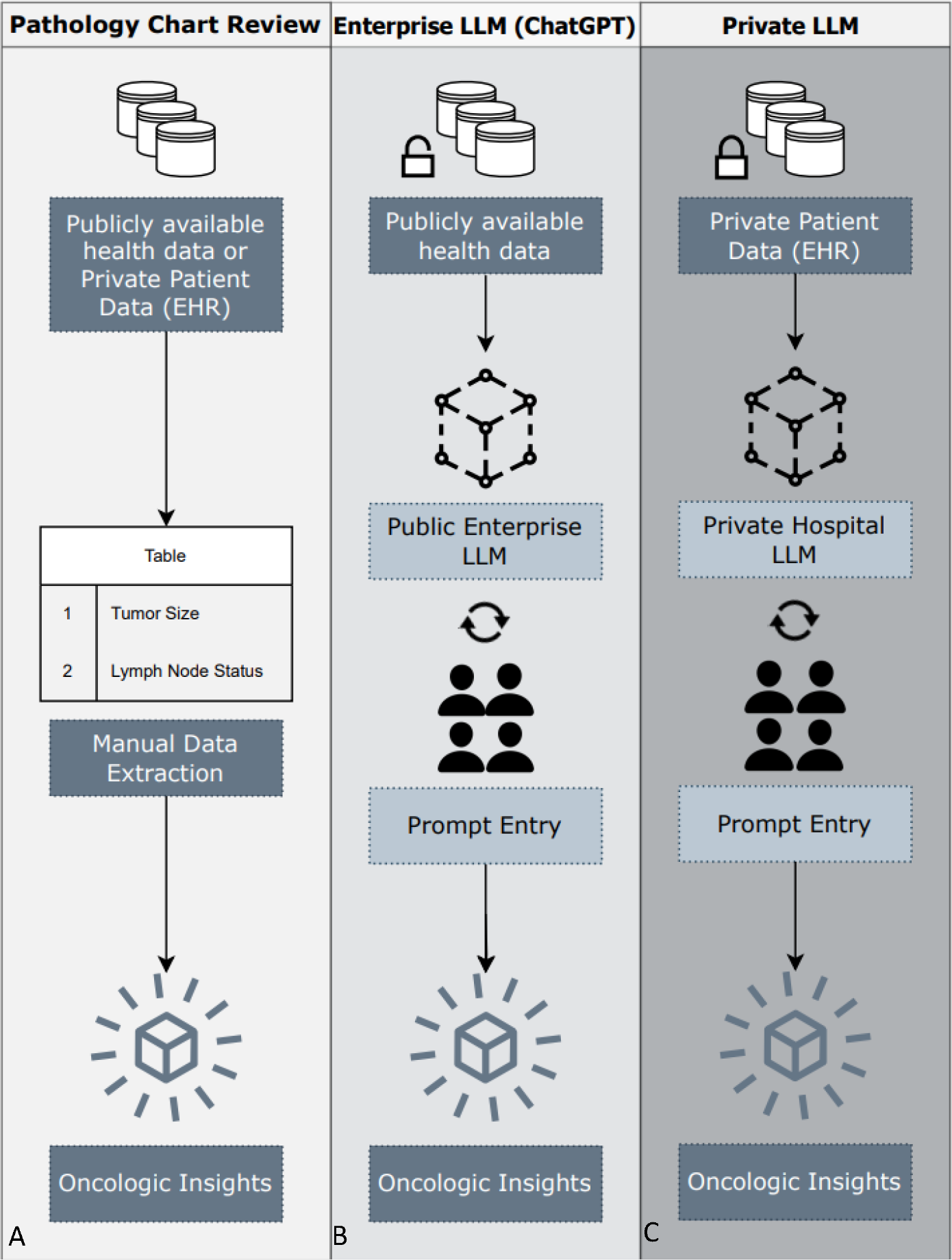
Overview of Medical Data Extraction Workflows: **(A) Pathology Chart Review:** Traditional approach of manual data extraction from publicly available databases or private electronic health records to obtain predetermined oncologic insights. **(B) Enterprise LLM (ChatGPT**): Due to regulatory constraints only publicly available data may be shared with enterprise LLMs. Prompt entry and question curation are used to gain oncologic insights. **(C) Private LLM**: Electronic health record data can be shared with a local hospital LLM and prompt entry with question curation can be used to gain oncologic insights.

## Methods

### Study Population

This study was approved by the Institutional Review Board of the Icahn School of Medicine at Mount Sinai. We queried our system wide database for a cohort of adult patients with diagnosis codes for thyroid cancer and who underwent thyroid surgery between 2010 and 2022. We reviewed 102 pathology reports from 102 patients and excluded reports if they were other organ site (n=10), benign (n=2), cytopathology (n=5), or outside review (n=1). We included 84 reports for analysis. Study flowchart is shown in **Figure 2**.

**Figure 2.**
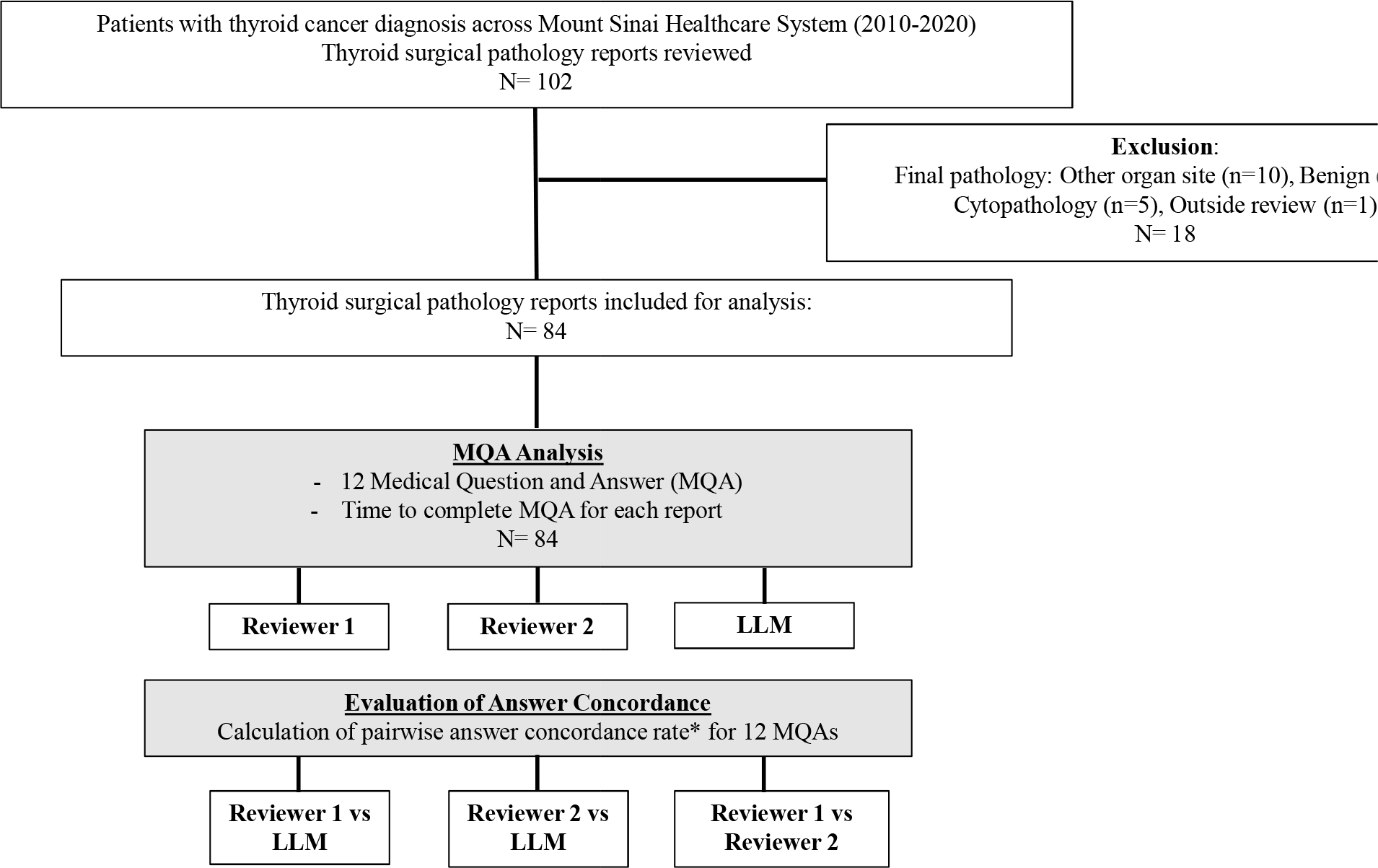
Flowchart of Study Design and Analysis. *Concordance rate: calculated as the total number of concordant answers/total number of answers for each of the 12 MQAs

### Development of Large Language Model

We used the publicly available, open-source FastChat-T5 3B-parameter LLM for our analysis. A limitation all LLMs have is the amount of context they may process at once. For reports of length greater than what the model could accommodate, we split report text into 1200 character long segments, followed by converting each of these segments into machine-readable numerical representations called embeddings. Since embeddings encode meaning, calculating similarity scores between segment embeddings and posed questions allowed us to retrieve the pieces of text most directly related to the content of the question. As such, three segments with the highest similarity scores were integrated to create the final context for the LLM. This context was made part of a plain language question for the LLM, alongside a question it was directed to answer.

### Development of Medical Question and Answering and Evaluation of Concordance

We formulated twelve questions with expert clinical input that extracted key information for the assessment of AJCC/TNM 8^th^ edition thyroid cancer staging and recurrence risk according to the American Thyroid Association Recurrence Risk Stratification System (5, 6). Two study authors (D.T.L and K.M) reviewed 84 thyroid surgical pathology reports and recorded answers to each of the twelve questions and time to complete answers for each report. Then we used the LLM to answer the same questions. For every question we determined if answers were concordant between: Reviewer 1 and the LLM, Reviewer 2 and the LLM, and between the two reviewers. Concordance rate for each pairwise comparison was then calculated as the total number of concordant answers divided by the total number of answers for each of the twelve questions (**Figure 2**). The average concordance rate and standard deviation of all questions were calculated for each pairwise comparison.

## Results

We report sample LLM responses and the concordance rates between reviewers and the LLM for each question in **Table 1**. 1,008 total questions were answered for 84 thyroid surgical pathology reports. Reviewers 1 and 2 were concordant at an overall rate of 99.1% (SD: 1.0%) with disagreement on 9 answers. Reviewers 1 and 2 took an average of 2.36 minutes and 1.48 minutes to respond to each pathology report and 206.9 minutes and 124.04 minutes for all reports, respectively. The LLM was concordant with reviewers 1 and 2 at an overall rate of 88.86% (SD: 7.02%) and 89.56 (SD: 7.20%). Average time to review each report for the LLM was 13.97 seconds/report and 19.56 minutes for all reports. The questions with the highest overall rates of concordant responses were questions requiring binary or categorical data extraction (*Is lymphatic invasion present, 100%, Is vascular invasion present, 98*.*81%, Where is the primary cancer located, 98*.*1%)*. The question with the lowest overall concordance was, *Were cervical lymph nodes present?* at 75%.

**Table 1.** 12 Medical Questions and Sample LLM Answers with Rates of Concordance Between the Reviewers and LLM. Abbreviations: LLM, Large language model; R1, Reviewer 1; R2, Reviewer 2

| Medical Question | Sample LLM Answer | Concordance (%) |  |  |  |
| --- | --- | --- | --- | --- | --- |
|  |  | R1, LLM | R2, LLM | R1, R2 | All 3 |
| **Staging** |  |  |  |  |  |
| **Tumor (T)** |  |  |  |  |  |
| *What is the size of the primary tumor?* | *The size of the primary tumor was 5.5 cm.* | 82.14 | 84.52 | 97.62 | 82.14 |
| *Does the tumor extend beyond the capsule of the thyroid?* | *No, the tumor does not extend beyond the capsule of the thyroid.* | 90.36 | 92.77 | 97.59 | 90.36 |
| *Where is the primary cancer located (right thyroid lobe, left thyroid lone or isthmus)?* | *The primary cancer is located in the right thyroid lobe.* | 98.81 | 100 | 98.81 | 98.81 |
| **Node (N)** |  |  |  |  |  |
| *Were any cervical lymph nodes present?* | *Yes, cervical lymph nodes were present.* | 76.19 | 75 | 98.81 | 75 |
| *If cervical lymph nodes are present, how many lymph nodes were positive for malignancy?* | *Three lymph node was positive for malignancy.* | 88.1 | 88.1 | 100 | 88.1 |
| **Stage (TNM)** |  |  |  |  |  |
| *What is the final pathology TNM stage of the thyroid cancer?* | *The final pathology TNM stage of the thyroid cancer is pT3a.* | 85.54 | 85.54 | 100 | 85.54 |
| **Recurrence Risk Factor** |  |  |  |  |  |
| *Is vascular invasion present?* | *Yes, vascular invasion is present.* | 98.81 | 98.81 | 100 | 98.81 |
| *Is lymphatic invasion present?* | *No, lymphatic invasion is not present.* | 100 | 100 | 100 | 100 |
| *What was the histology of the primary cancer?* | *The histology of the primary thyroid cancer was papillary carcinoma.* | 86.9 | 86.9 | 100 | 86.9 |
| *Was there a variant of thyroid cancer such as tall cell, hobnail variant, or columnar cell variant present indicating more aggressive histology?* | *No, a variant of thyroid cancer such as tall cell, hobnail variant, or columnar cell variant was not present indicating more aggressive histology.* | 89.29 | 91.67 | 97.62 | 89.29 |
| *Was there a second thyroid cancer present?* | *No, there was no second thyroid cancer present.* | 82.14 | 83.33 | 98.81 | 82.14 |
| *If there was a second thyroid cancer present, what was the histology?* | *The histology of the second thyroid cancer is classical variant papillary thyroid carcinoma.* | 88.1 | 88.1 | 100 | 88.1 |
| **Overall Questions, (SD)** |  | 88.86 (7.02) | 89.56 (7.20) | 99.1 (1.0) | 88.76 (7.2) |

## Discussion

We demonstrate and evaluate the extraction performance of a privacy-preserving LLM for a specific clinical NLP task. The LLM took 19.56 minutes to evaluate and respond to all pathology reports, whereas it required an additional 187 minutes and 105 minutes for the reviewers to complete the same task— demonstrating a considerable reduction in time. Regarding accuracy of responses, we find that rates of response concordance were higher amongst questions tasked with simpler binary or categorical responses. Increase in task complexity requiring textual interpretation and inconsistent word prompting such as asking whether there was “cervical” lymph nodes present resulted in the lowest rate of concordance. Furthermore, the question of size of the primary tumor also seemed to be relatively straightforward but only had an overall concordance rate of 82%.

The augmentation of poorer performing MQA may lie in the improvement of prompt engineering— an emerging subfield where domain specific knowledge and linguistics are optimized to design questions that yield the best performing response to a task, in addition to more expressive embeddings that better help localize relevant text (7). For example, “cervical” does not appear in most pathological reports verbatim, possibly limiting the model’s ability to respond appropriately to the question regarding the presence of cervical lymph nodes. Also, the LLM often incorrectly identified the size of the “primary tumor” and would instead provide a dimension from another specimen in the report, such as the overall thyroid lobe. This response accuracy may also be improved by modifying the question prompt and will be the focus of future work.

Overall, the use of our current methodology is an advance from prior NLP efforts with limitations such as restrictive data preprocessing and the inability to handle multiple positive diagnoses (8-10). Our method of developing and deploying LLMs behind a healthcare institution’s own computing resources, ensures that centers could utilize this emerging technology while maintaining patient privacy. Additionally, since we utilize the inherent reasoning ability of such models, they do not require any task specific fine-tuning, and by extension can be operated inexpensively. Furthermore, the increased language capacity of latest generation of LLMs allows for institutions to deploy their own data for in-context learning only while achieving a reasonable performance.

## Conclusion

We envision that LLMs will allow medical institutions to harness cutting-edge advances in NLP for timely and privacy-preserving MQA data extraction from pathology reports and other clinical narratives for the provision of real-time, essential oncologic insights.

## Data Availability

All data produced in the present study are available upon reasonable request to the authors.

## References

1. Burger G, Abu-Hanna A, de Keizer N, Cornet R. Natural language processing in pathology: a scoping review. J Clin Pathol. 2016.

2. Yim WW, Yetisgen M, Harris WP, Kwan SW. Natural Language Processing in Oncology: A Review. JAMA Oncol. 2016;2(6):797–804.

3. Thirunavukarasu AJ, Ting DSJ, Elangovan K, Gutierrez L, Tan TF, Ting DSW. Large language models in medicine. Nat Med. 2023;29(8):1930–40.

4. Yang X, Chen A, PourNejatian N, Shin HC, Smith KE, Parisien C, et al. A large language model for electronic health records. npj Digital Medicine. 2022;5(1):194.

5. Tuttle RM, Haugen B, Perrier ND. Updated American Joint Committee on Cancer/Tumor-Node-Metastasis Staging System for Differentiated and Anaplastic Thyroid Cancer (Eighth Edition): What Changed and Why? Thyroid. 2017;27(6):751–6.

6. Haugen BR, Alexander EK, Bible KC, Doherty GM, Mandel SJ, Nikiforov YE, et al. 2015 American Thyroid Association Management Guidelines for Adult Patients with Thyroid Nodules and Differentiated Thyroid Cancer: The American Thyroid Association Guidelines Task Force on Thyroid Nodules and Differentiated Thyroid Cancer. Thyroid. 2016;26(1):1–133.

7. Savova GK, Danciu I, Alamudun F, Miller T, Lin C, Bitterman DS, et al. Use of Natural Language Processing to Extract Clinical Cancer Phenotypes from Electronic Medical Records. Cancer Res. 2019;79(21):5463–70.

8. Datta S, Bernstam EV, Roberts K. A frame semantic overview of NLP-based information extraction for cancer-related EHR notes. J Biomed Inform. 2019;100:103301.

9. Schadow G, McDonald CJ. Extracting structured information from free text pathology reports. AMIA Annu Symp Proc. 2003;2003:584–8.

10. Mitchell JR, Szepietowski P, Howard R, Reisman P, Jones JD, Lewis P, et al. A Question-and-Answer System to Extract Data From Free-Text Oncological Pathology Reports (CancerBERT Network): Development Study. J Med Internet Res. 2022;24(3):e27210.

